# Machine learning identifies shared blood transcriptional biomarkers and immune correlates across antiphospholipid syndrome and systemic sclerosis

**DOI:** 10.64898/2026.01.20.26344459

**Authors:** Ren-Ju Yang, Jin-Chan Liu, Ya-Wen Liu, Fei He

## Abstract

Antiphospholipid syndrome (APS) and systemic sclerosis (SSc) are immune-mediated multisystem autoimmune diseases with distinct clinical phenotypes but overlapping pathogenic themes, including immune dysregulation, chronic inflammation, and endothelial injury. Using peripheral blood transcriptome datasets from the Gene Expression Omnibus (GSE102215: 9 APS/9 controls; GSE231691: 49 SSc/18 controls), we performed differential expression analysis within each cohort (limma; |log2FC|>1, P<0.05) and identified 281 genes dysregulated in the same direction in both diseases (100 upregulated and 181 downregulated). Enrichment analyses highlighted interferon-related and cytokine/inflammatory signaling programs in APS and SSc. To derive a compact diagnostic signature, we combined random forest feature ranking with a single-hidden-layer artificial neural network, prioritizing five shared candidate biomarkers (S100A8, IER5L-AS1, LTK, PRR5-ARHGAP8, and PCDH1). Each gene showed consistent case-control differences in both cohorts (P<0.001) and achieved good discrimination (AUC>0.75), with S100A8 performing most consistently (AUC=0.98 in APS; AUC=0.88 in SSc). CIBERSORT deconvolution indicated a myeloid-skewed blood profile in both diseases, characterized by higher neutrophil and monocyte/macrophage signals; SSc additionally showed stronger inferred CD4+ T cell and NK cell signals. S100A8 expression correlated with inferred neutrophil abundance in both cohorts (APS r=0.62; SSc r=0.58; P<0.05). Finally, miRNA-target prediction and DSigDB drug-signature enrichment generated regulatory and pharmacologic hypotheses, including immune-regulatory miRNAs (e.g., miR-155 and miR-146a) and candidate compounds (celecoxib, tamibarotene, HMN-176, and XMD14-99). Overall, these results nominate shared blood transcriptional markers and immune correlates across APS and SSc for follow-up validation.

## Introduction

Autoimmune diseases arise from loss of immune tolerance and can cause chronic inflammation and multisystem tissue injury. APS is defined by antiphospholipid antibody-associated thrombosis and pregnancy morbidity [1], whereas SSc is characterized by immune activation, microvascular damage, and progressive fibrosis, with classification refined through the 2013 ACR/EULAR criteria [2,3]. Despite distinct clinical phenotypes, both diseases involve perturbed innate and adaptive immune responses, altered cytokine networks, and endothelial dysfunction [4,5]. These shared mechanisms raise the possibility of convergent blood transcriptional programs that could help explain clinical overlap and support cross-disease biomarker discovery [6,7].

Large public repositories such as the Gene Expression Omnibus (GEO) make it possible to compare disease-associated transcriptional signatures across independent cohorts [8,9]. Cross-disease analyses have repeatedly shown convergence on conserved inflammatory modules, particularly interferon-driven programs, alongside disease-specific components [10]. In parallel, machine-learning methods are increasingly used to reduce high-dimensional expression profiles to smaller feature sets with potential diagnostic or stratification value [11]. Random forests provide nonlinear feature ranking that is relatively robust to correlated predictors [12], whereas neural networks can capture interactions among selected features and offer a complementary view of feature importance [13]. Using both approaches can help prioritize signals that are less likely to be driven by a single cohort’s idiosyncrasies [14].

Bulk whole-blood transcriptomes are strongly influenced by immune-cell composition. Neutrophil extracellular trap formation and monocyte activation have been implicated in APS [15], while macrophage polarization and T-cell subset imbalance contribute to SSc pathogenesis [5]. However, shared versus disease-specific immune-cell patterns across APS and SSc have not been systematically compared under a consistent analytic framework, and the relationship between candidate biomarker genes and inferred immune-cell fractions remains insufficiently characterized. Because bulk signatures can reflect shifts in cell proportions, activation states, or both, integrating digital cytometry with gene-level analyses can clarify the biological basis of candidate markers.

In this study, we integrated peripheral blood transcriptome datasets for APS and SSc to identify concordantly dysregulated genes, prioritized a small set of shared biomarkers using complementary machine-learning models, and estimated immune-cell composition by digital cytometry. We then examined gene–immune associations, predicted potential upstream miRNAs, and performed drug-signature enrichment to generate testable regulatory and translational hypotheses [16,17].

## Materials and methods

### Data sources and preprocessing

Two peripheral blood transcriptome datasets were downloaded from GEO: GSE102215 (APS; 9 patients and 9 healthy controls; platform GPL16791) and GSE231691 (SSc; 49 patients and 18 healthy controls; platform GPL20301). All samples were derived from Homo sapiens peripheral blood (Table 1).

**Table 1.**
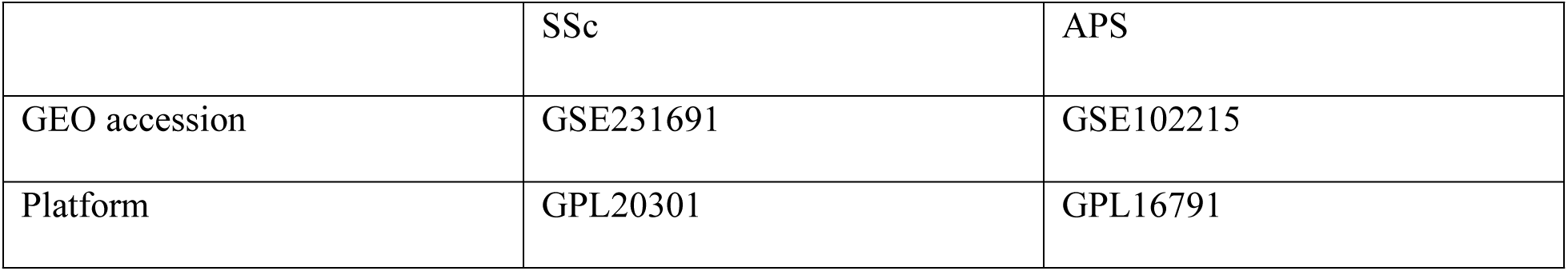
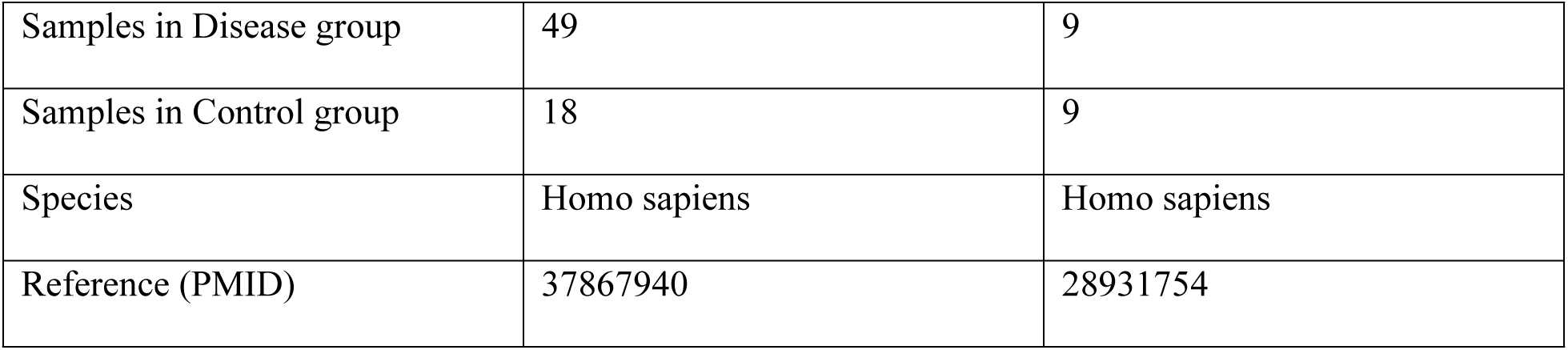
Summary of datasets used in this study.

RNA-seq data from GEO datasets GSE102215 and GSE231691 were analyzed. Gene-level raw count matrices were extracted and sample grouping information was constructed. Size-factor normalization was performed using DESeq2. Differential expression analysis was conducted based on a negative binomial model with multiple testing correction using the Benjamini–Hochberg method. Genes with padj < 0.05 and |log2FoldChange| > 1 were considered differentially expressed. Variance stabilizing transformation (VST) was applied for visualization analyses.

Because the datasets were generated on different platforms and represent independent studies, we did not merge expression matrices across cohorts. Instead, differential expression and feature ranking were performed within each dataset, and cross-disease signals were defined by concordant results observed in both datasets. This design limits direct platform confounding while enabling comparison at the level of shared biological signals [19].

### Differential expression and shared gene definition

Differential expression was assessed separately for each dataset using limma [20]. Genes with |log2FC| > 1 and nominal P < 0.05 were considered differentially expressed, with log2FC > 1 indicating upregulation and log2FC < -1 indicating downregulation. Given the exploratory aim and the small APS cohort, we report nominal P values for DEG definition and interpret downstream results with appropriate caution.

Multiple testing and validation strategy. Because genome-wide differential-expression screening involves thousands of simultaneous tests, we interpreted nominal P values from limma as exploratory signals and emphasized robustness through cross-dataset concordance: candidates were prioritized only when they showed consistent direction and met the DEG criteria in independent cohorts. For enrichment analyses, we relied on the multiple-testing–adjusted P values returned by clusterProfiler (Benjamini–Hochberg procedure) and used adjusted P values (or FDR q values for GSEA) to determine significance. For diagnostic evaluation, ROC/AUC estimates reflect within-cohort discrimination and are reported as preliminary in the absence of an independent APS validation cohort.

Shared differentially expressed genes (shared DEGs) were defined as genes that met the DEG criteria in both datasets and changed in the same direction. Overlap was visualized with Venn diagrams, and volcano plots and heatmaps were used to summarize expression patterns across samples.

### Functional enrichment analyses

Gene set enrichment analysis (GSEA) was performed for each dataset using clusterProfiler with Hallmark gene sets (category H) from msigdbr [18]. Genes were ranked by log2FC, and enrichment results were visualized with enrichplot (e.g., enrichment curves, bubble plots, and ridge plots).

To interpret shared DEGs, Gene Ontology (GO) and Kyoto Encyclopedia of Genes and Genomes (KEGG) enrichment analyses were performed using clusterProfiler with org.Hs.eg.db for identifier conversion [18]. GO enrichment covered biological process, cellular component, and molecular function categories, and KEGG enrichment was performed for Homo sapiens (hsa).

### Machine-learning feature selection and diagnostic evaluation

Shared DEGs were used as candidate features for biomarker prioritization. Random forest models were trained separately in the APS and SSc datasets using randomForest with 500 trees, with disease status (patient=1, control=0) as the response [12]. Feature importance was quantified by mean decrease in Gini [21], and the top 30 genes were extracted from each dataset. The intersection of these top-ranked genes across diseases was taken as the set of shared candidate biomarkers.

Artificial neural network (ANN) classifiers were then trained using nnet with z-score-normalized expression values of the shared candidate biomarkers [13]. Models used a single hidden layer (3 neurons), a maximum of 500 iterations, a learning rate of 0.01, and a fixed random seed (123). Gene importance was summarized from network weights, and model structure was visualized using NeuralNetTools.

Diagnostic performance of each biomarker was evaluated by receiver operating characteristic (ROC) analysis using pROC, reporting AUC and 95% confidence intervals [22]. Given the limited APS sample size, these performance estimates reflect within-cohort discrimination and should not be interpreted as external validation.

### Immune deconvolution and gene–immune association

Immune-cell composition was inferred using CIBERSORT to estimate the relative fractions of 22 immune cell types (LM22) [23]. Permutations were set to 1000, and samples with CIBERSORT output P > 0.05 were excluded.

Differences in immune-cell fractions between disease and control groups were assessed using Wilcoxon rank-sum tests and visualized with stacked bar charts and boxplots. Associations between biomarker expression and immune-cell fractions were evaluated using Spearman rank correlation [22].

### miRNA prediction and drug-signature enrichment

Potential upstream miRNAs targeting the prioritized biomarkers were retrieved from miRWalk [24]. An miRNA–mRNA interaction network was constructed and visualized in Cytoscape [25]. miRNAs with established roles in immune regulation (e.g., miR-155 and miR-146a) were highlighted based on prior evidence in autoimmune settings [26].

Candidate compounds were nominated via drug–gene signature enrichment using the Drug Signature Database (DSigDB) [27]. DSigDB integrates drug-induced gene expression signatures, enabling identification of compounds whose signatures overlap with the genes of interest [28]. Drugs were ranked by enrichment statistics (including adjusted P values and combined scores), and the top candidates are reported as hypothesis-generating leads.

### Software and statistical analysis

Unless otherwise specified, analyses were conducted in R (v4.4.1) [18]. Two-sided P < 0.05 was considered statistically significant. Where multiple testing correction was applied (e.g., enrichment analyses), false discovery rate (FDR)-adjusted values reported by the corresponding methods were used.

## Results

### Study overview

We retrieved public APS and SSc peripheral blood transcriptome datasets from GEO and applied a within-dataset workflow for preprocessing and differential expression. We then identified concordantly dysregulated genes shared between diseases, characterized these genes with enrichment analyses, prioritized shared diagnostic candidates using machine learning, estimated immune-cell composition by digital cytometry, and explored gene–immune correlations. Finally, we generated regulatory (miRNA) and pharmacologic (drug-signature) hypotheses (Fig 1).

**Fig 1.**
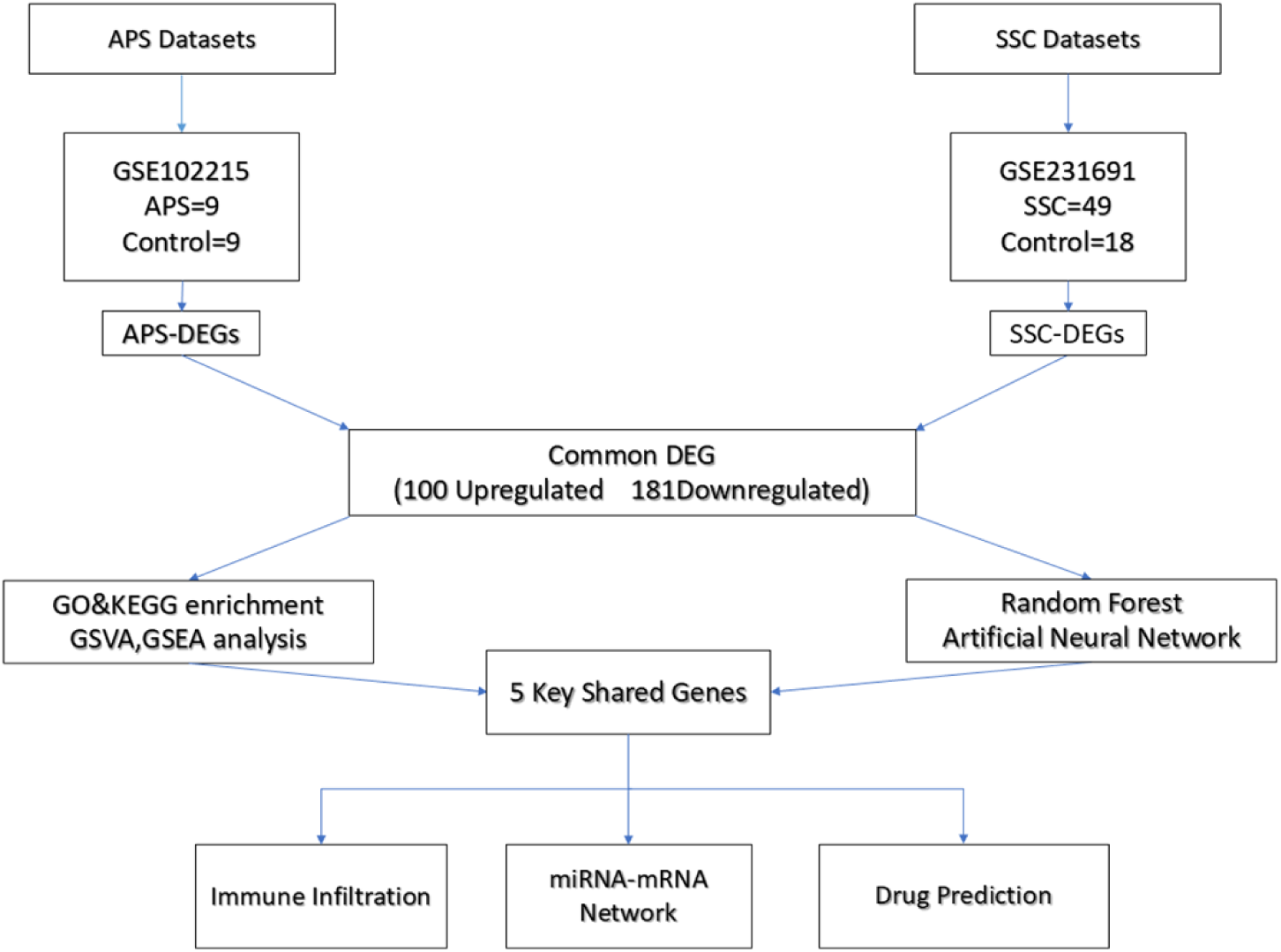
Study design and analytical workflow used to identify shared biomarkers and immune correlates across APS and SSc.

### Shared differentially expressed genes across APS and SSc

In the APS dataset, limma identified 2,744 upregulated and 1,058 downregulated genes using |log2FC| > 1 and P < 0.05. In the SSc dataset, 361 genes were upregulated and 1,566 were downregulated (1,927 DEGs total). Intersecting concordant results yielded 281 shared DEGs (100 commonly upregulated and 181 commonly downregulated; Fig 2; S1–S2 Tables). The larger number of DEGs in the APS cohort may reflect the small sample size, inter-individual variability, and the use of nominal P values without additional filtering [29,30].

**Fig 2.**
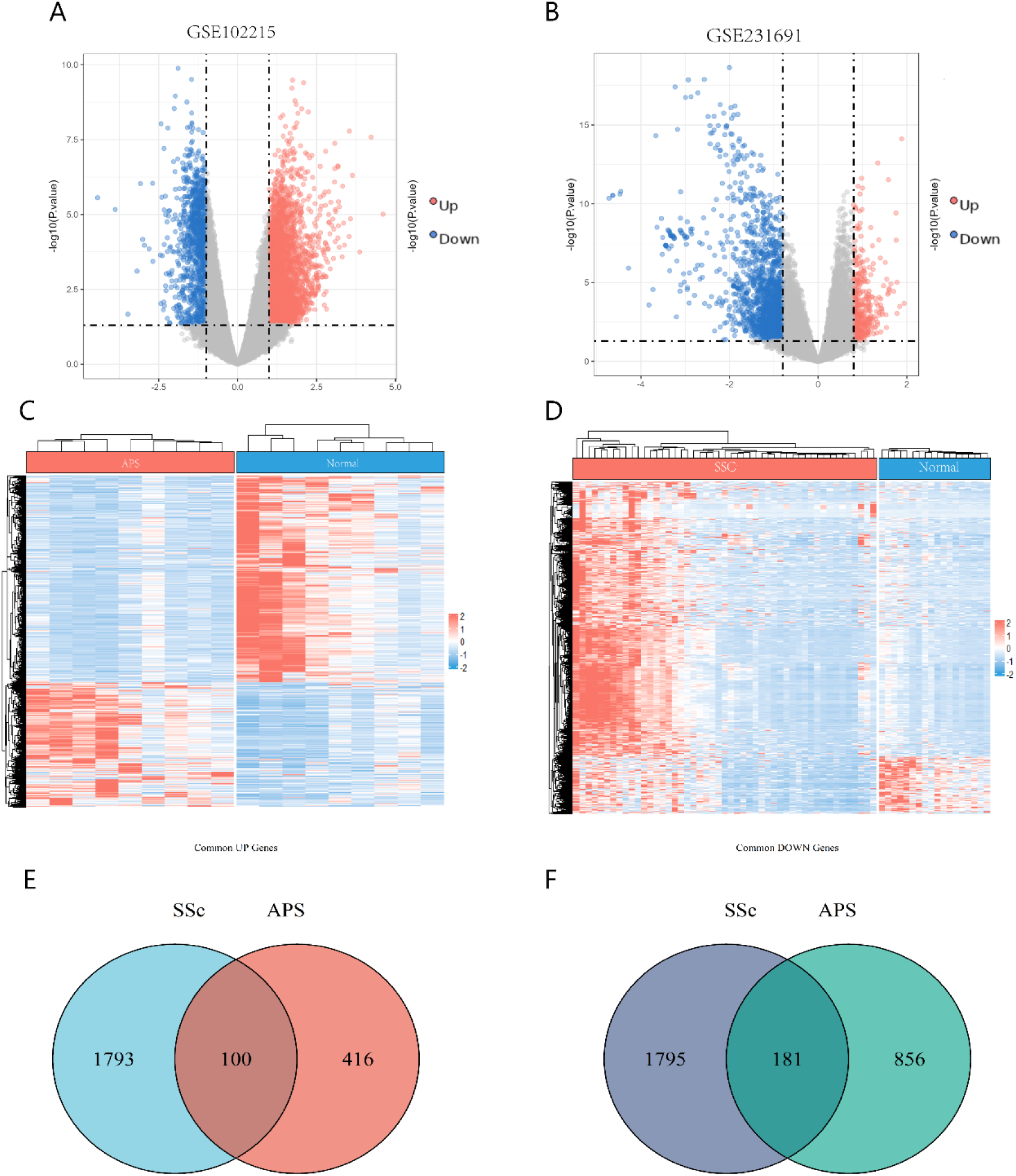
Differential expression and shared DEGs in APS and SSc. Volcano plots, expression heatmaps, and clustering summarize differential expression in each cohort and support the 281 concordantly regulated shared DEGs (100 up, 181 down). (A–C–E) APS: (A) volcano plot; (C) DEG heatmap; (E) hierarchical clustering with group annotation. (B–D–F) SSc: (B) volcano plot; (D) DEG heatmap; (F) hierarchical clustering with group annotation.

The figures further illustrate these shared signals. Volcano plots (Fig 2A for APS; Fig 2B for SSc) summarize differential expression between cases and controls. Heatmaps (Fig 2C–D) show the expression patterns of shared DEGs across samples, and hierarchical clustering (Fig 2E–F) separates disease and control samples in both datasets. Together, these visualizations support that the shared gene set captures reproducible case–control differences in each cohort.

### Pathway enrichment reveals convergent immune and inflammatory programs

Hallmark GSEA highlighted immune and inflammatory pathways in both diseases. In APS, interferon alpha and interferon gamma response signatures were prominent, along with other cytokine- and inflammation-related programs (Fig 3A–B). SSc showed a broadly similar pattern (Fig 3C–D). These findings align with prior reports implicating interferon and cytokine dysregulation in APS and SSc [5,15].

**Fig 3.**
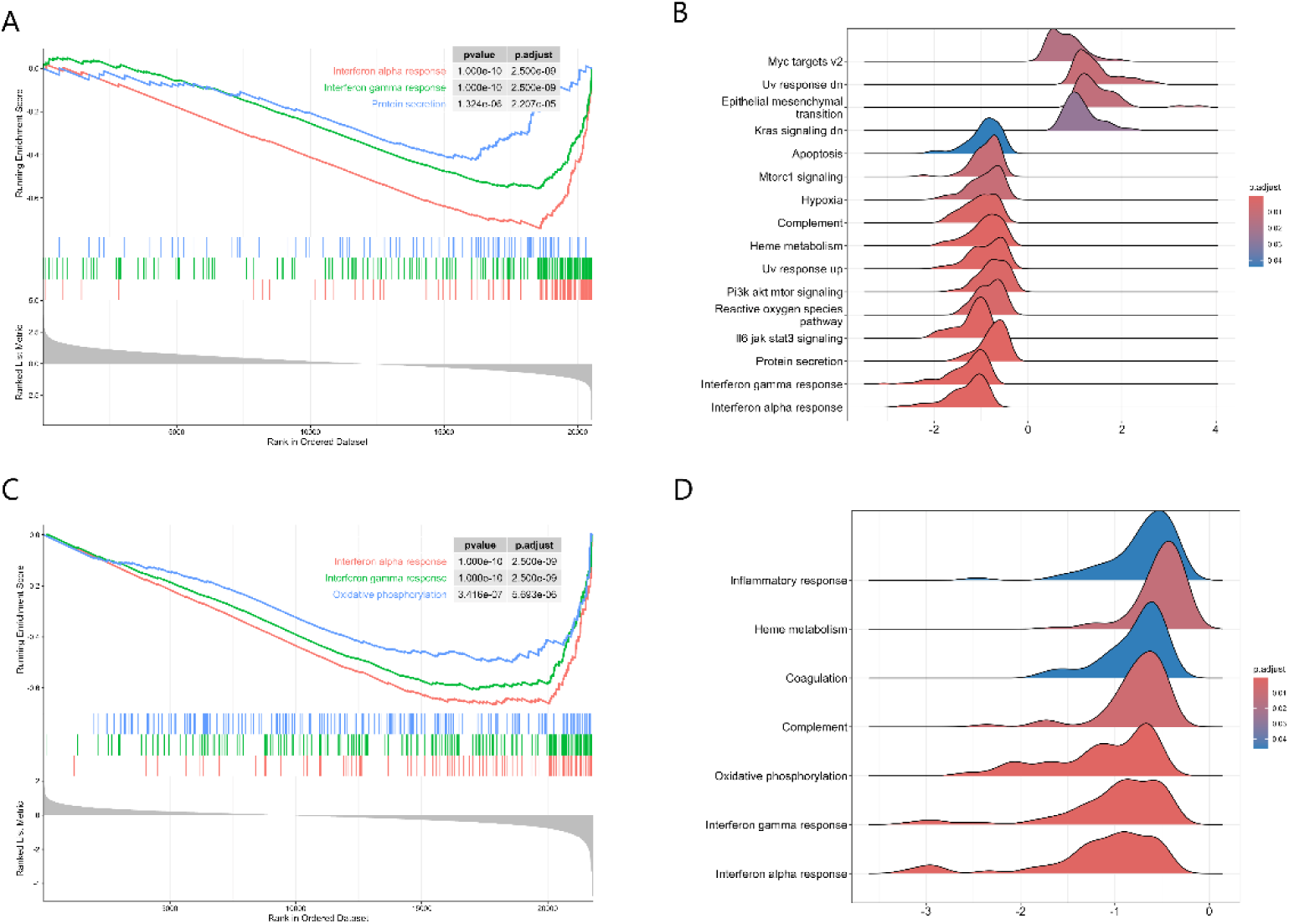
Hallmark GSEA in APS and SSc. GSEA highlights immune/inflammatory programs shared across diseases. APS: (A) enrichment curves for representative Hallmark sets; (B) ridge plot summarizing NES across Hallmark sets (colored by FDR). SSc: (C) enrichment curves for representative Hallmark sets; (D) ridge plot summarizing NES across Hallmark sets (colored by FDR).

GO and KEGG enrichment analyses of shared DEGs also pointed to immune activation and inflammatory signaling (Fig 4). GO terms in both cohorts were dominated by immune response and host-defense functions (Fig 4A–B), while KEGG results included pathways such as TNF and NF-kB signaling (Fig 4C–D), consistent with their roles in chronic inflammation [32].

**Fig 4.**
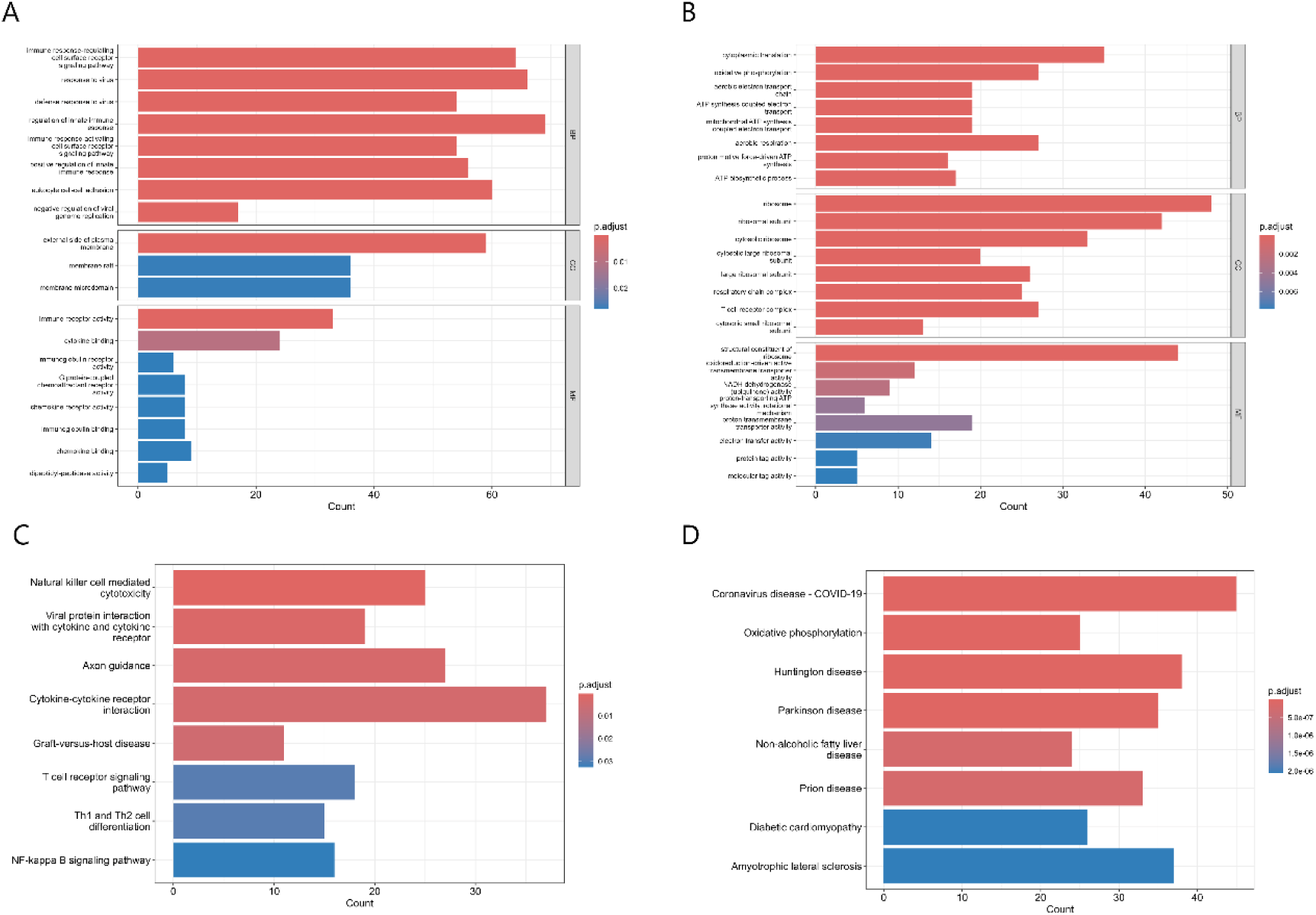
GO and KEGG enrichment of shared DEGs. GO and KEGG analyses of shared DEGs implicate immune and inflammatory functions. GO bar plots: APS (A) and SSc (B), grouped by BP/CC/MF. KEGG bar plots: APS (C) and SSc (D), highlighting significantly enriched pathways.

### Machine learning prioritizes five shared blood biomarkers

Random forest models trained in each cohort (500 trees) ranked genes by importance. The top 30 features from SSc (Fig 5A) and APS (Fig 5B) were intersected (Fig 5C), yielding five shared high-importance genes: S100A8, IER5L-AS1, LTK, PRR5-ARHGAP8, and PCDH1. Because these genes were selected independently in two datasets, they represent robust cross-disease candidates [11,13].

**Fig 5.**
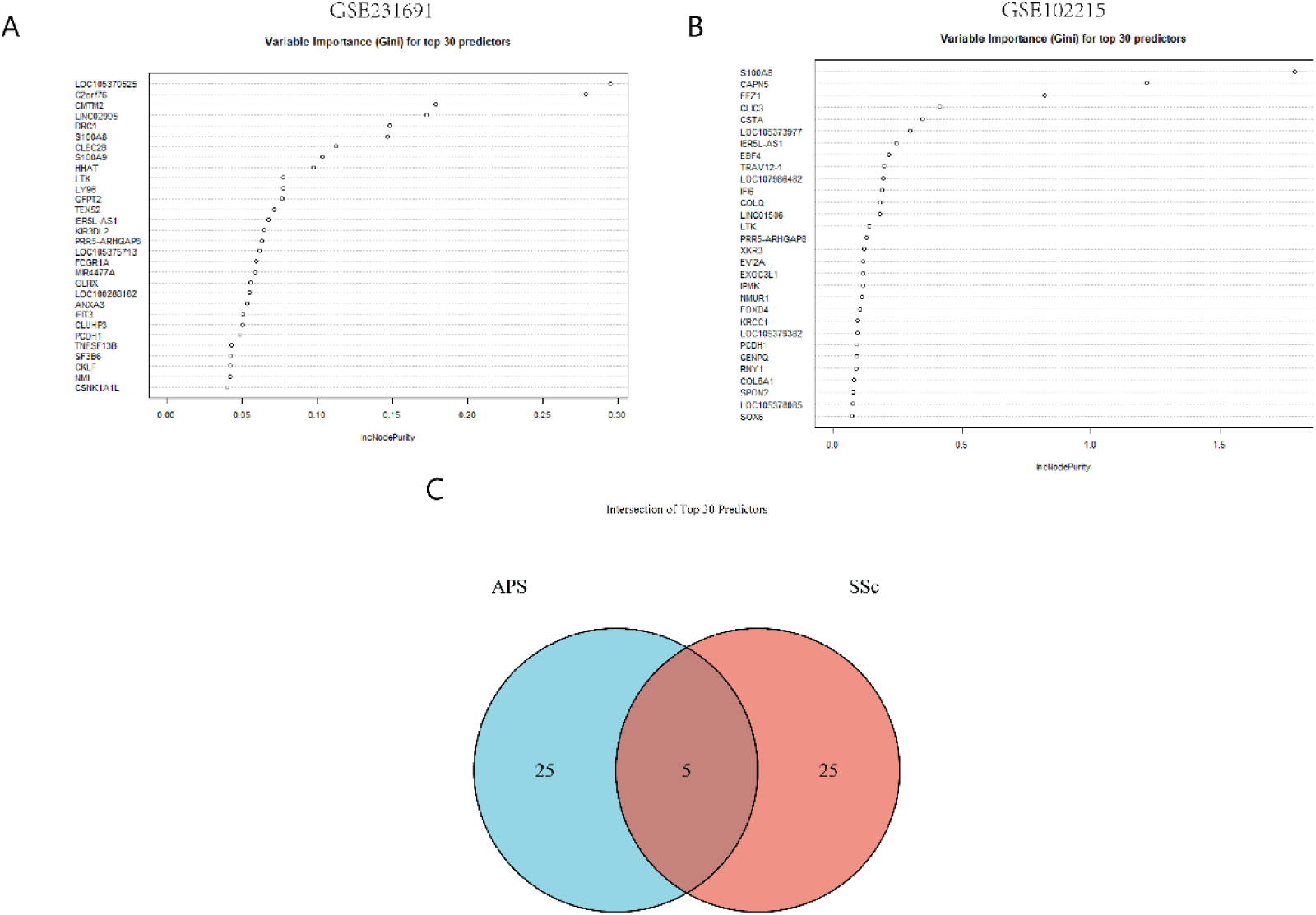
Random forest feature ranking and overlap across cohorts. Random forest importance ranks the top predictive genes in SSc (A) and APS (B) (IncNodePurity). (C) Venn overlap of the top 30 genes identifies five shared high-importance candidates: S100A8, IER5L-AS1, LTK, PRR5-ARHGAP8, PCDH1.

ROC analyses indicated good within-cohort discrimination for each gene in both datasets (AUC > 0.75) [22]. In APS, S100A8 achieved an AUC of 0.98 (95% CI 0.78–0.99), and the remaining genes also showed high AUC values (IER5L-AS1 0.96; LTK 0.93; PRR5-ARHGAP8 0.95; PCDH1 0.92; Fig 6A–E). In SSc, S100A8 achieved an AUC of 0.88 (95% CI 0.79–0.95), and the remaining genes had AUC values > 0.72 (Fig 6F–J). Notably, S100A8 showed consistently strong performance across cohorts.

**Fig 6.**
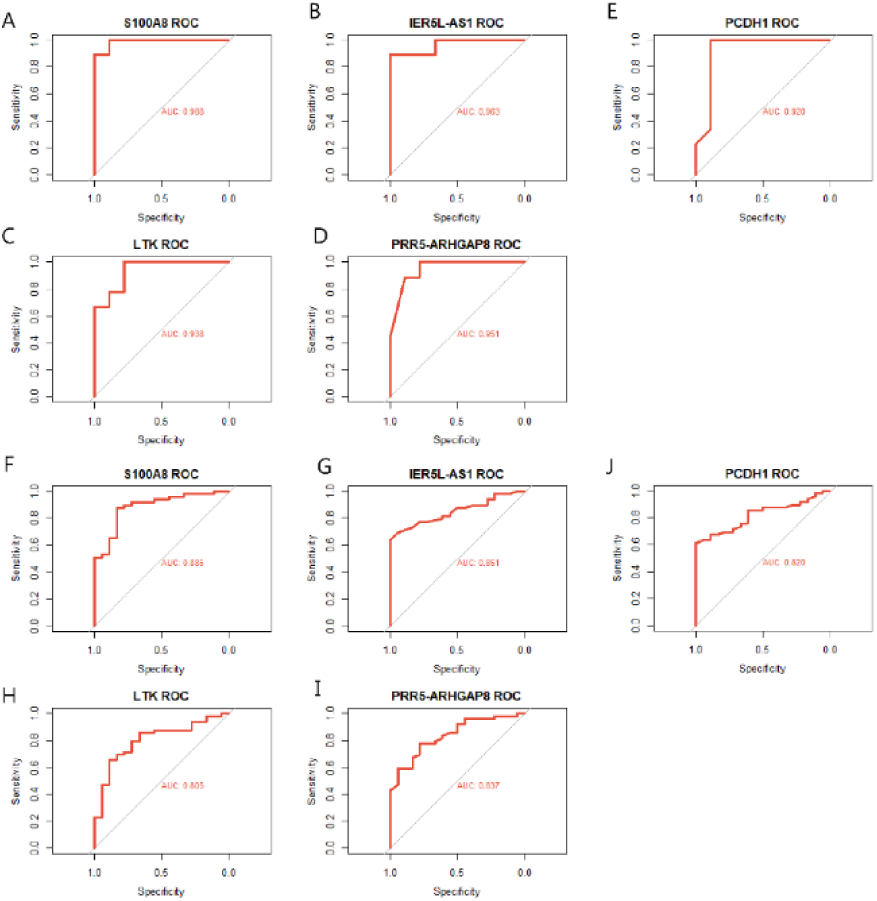
ROC performance of five shared biomarkers in APS and SSc. ROC curves for each biomarker in APS (A–E) and SSc (F–J): S100A8, IER5L-AS1, PCDH1, LTK, and PRR5-ARHGAP8 (AUC and 95% CI shown on plots). All genes achieve AUC > 0.75, supporting cross-disease discriminative potential.

ANN classifiers trained on the five-gene set provided complementary evidence that these markers capture disease-associated expression patterns. Model structures for APS and SSc are shown in Fig 7A–B. Boxplots of z-score–normalized expression (Fig 7C–D) show significant case–control differences for all five genes (Wilcoxon rank-sum test, P < 0.001), with concordant direction across APS and SSc.

**Fig 7.**
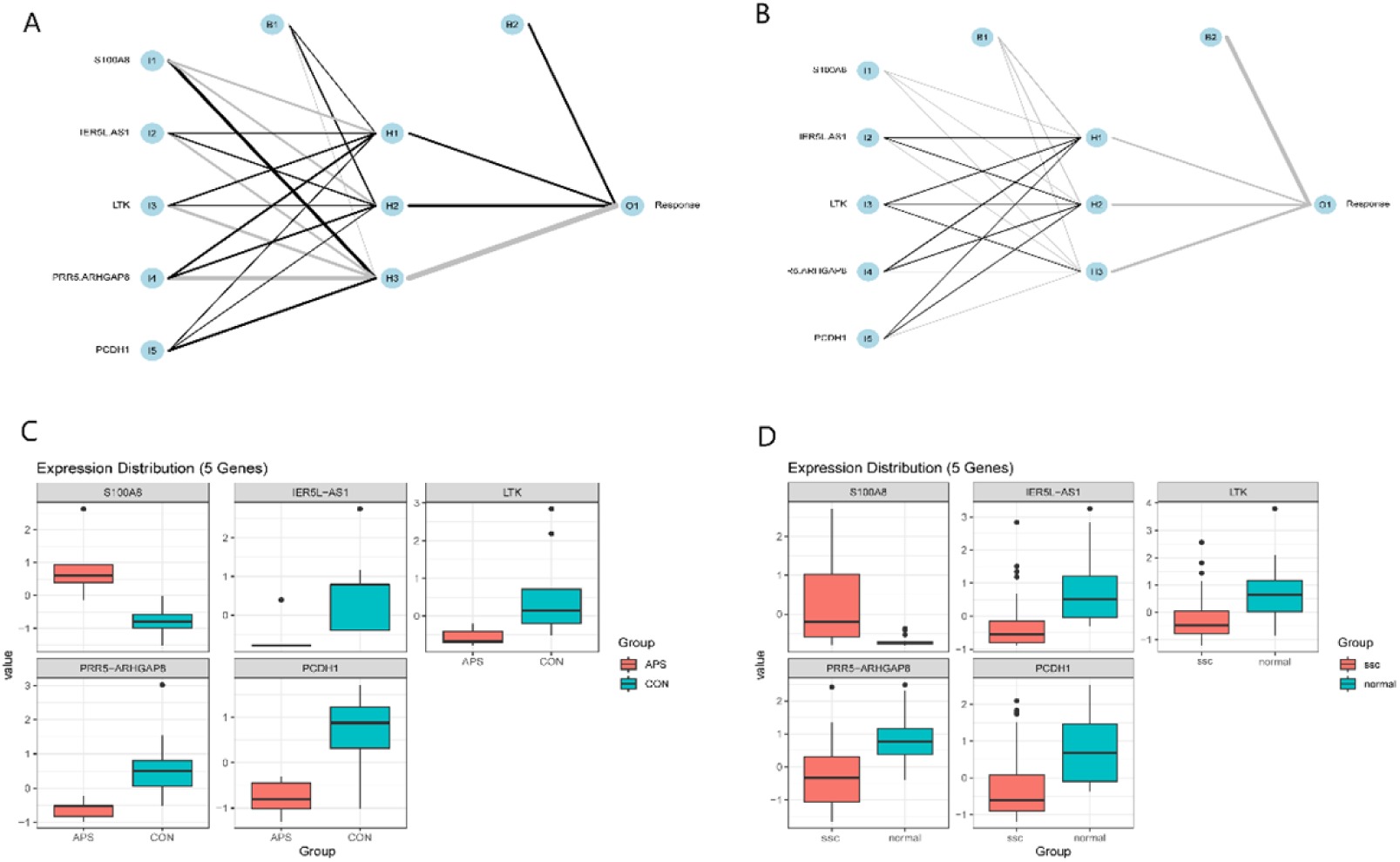
ANN modeling and expression validation of five shared genes. ANN models using the five genes as inputs in APS (A) and SSc (B) (single hidden layer with 3 neurons; max 500 iterations; learning rate 0.01; seed 123). Boxplots show Z-score–normalized expression differences between cases and controls in APS (C) and SSc (D) (Wilcoxon rank-sum test, P < 0.001).

### Immune infiltration patterns and gene–immune associations

CIBERSORT deconvolution indicated a myeloid-dominant immune profile in both diseases. Immune-cell fraction heatmaps (Fig 8A for APS; Fig 8C for SSc) show elevated myeloid-associated signals, and clustering based on inferred immune-cell profiles separated cases from controls in both cohorts (Fig 8B and Fig 8D) [23]. In SSc, inferred CD4+ T-cell and NK-cell signals were more pronounced than in APS (Fig 8C), consistent with broader adaptive immune involvement reported in SSc [4,5].

**Fig 8.**
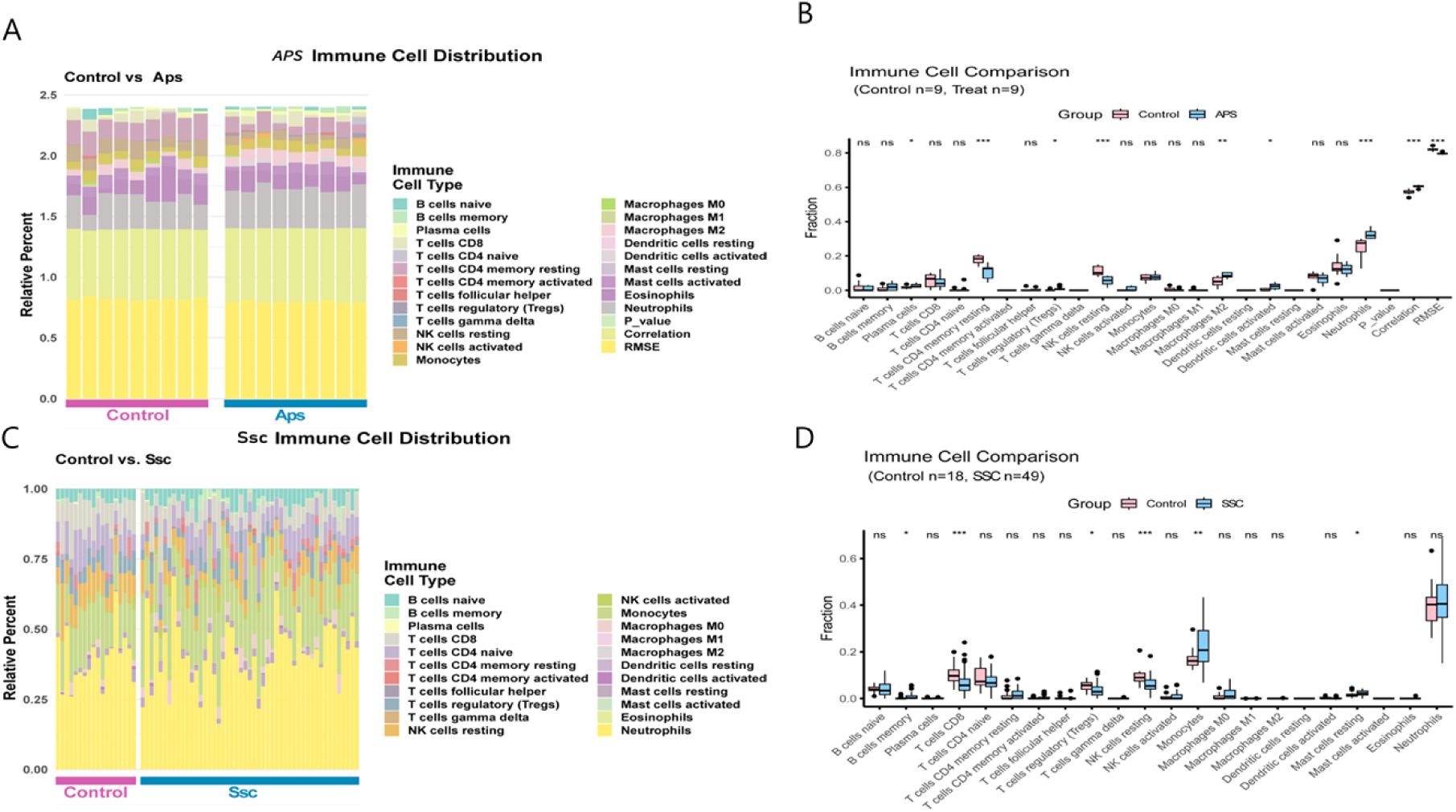
Immune cell infiltration patterns in APS and SSc. CIBERSORT-estimated immune-cell fractions across samples. APS: (A) heatmap of 22 immune cell types by sample; (B) clustering with group annotation. SSc: (C) heatmap; (D) clustering with group annotation.

Correlation analyses linked biomarker expression to inferred immune-cell fractions (Fig 9). S100A8 expression correlated positively with neutrophil abundance in both datasets (APS r = 0.62; SSc r = 0.58; P < 0.05), consistent with its known myeloid expression and role in inflammation [31,32]. PRR5-ARHGAP8 and PCDH1 showed strong correlations with resting NK cells across both diseases (|r| ≥ 0.6; P < 0.001), suggesting that the selected biomarkers reflect variation in both myeloid and lymphoid compartments (Fig 9; S3–S4 Tables).

**Fig 9.**
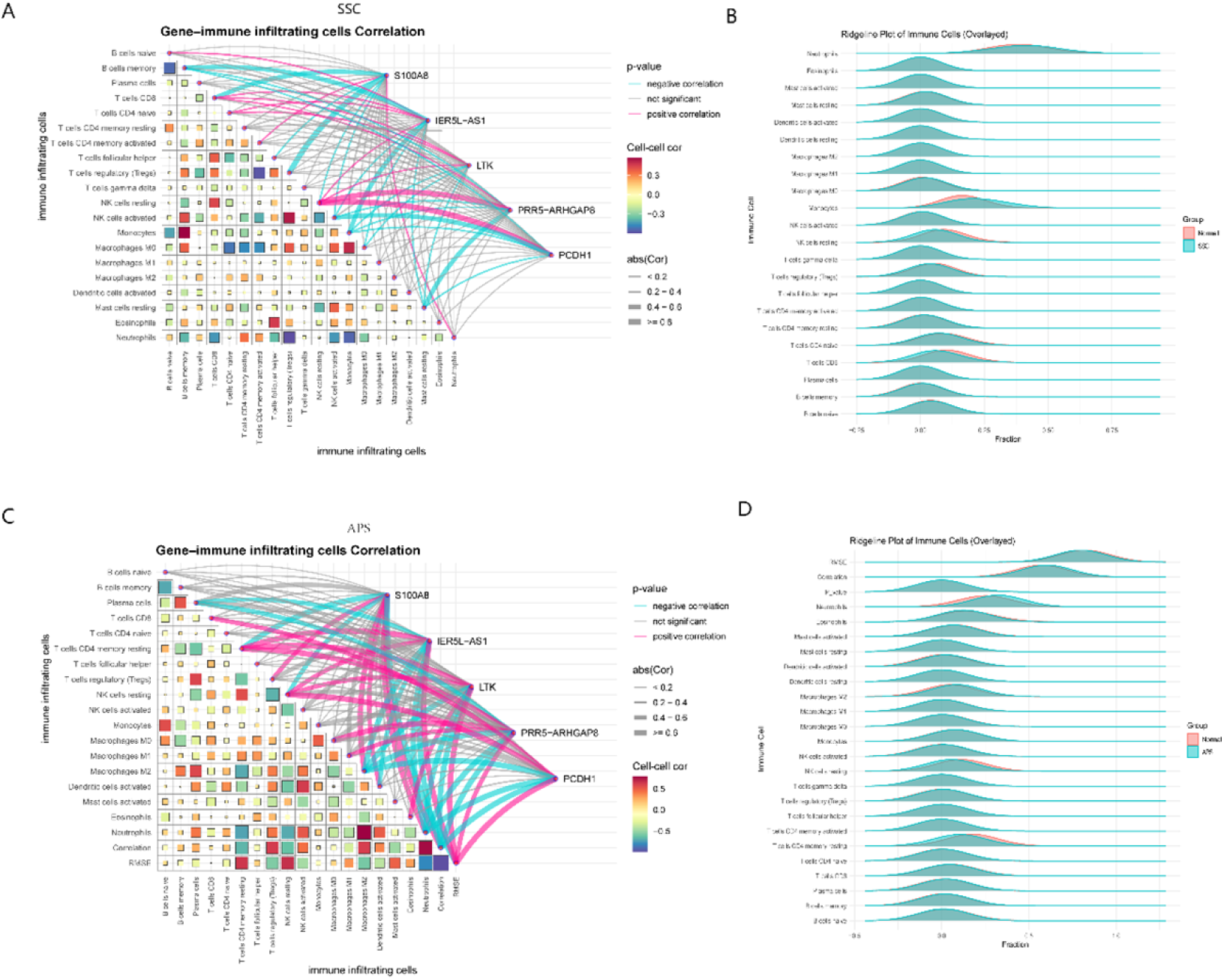
Gene–immune associations in APS and SSc. Spearman associations between five biomarkers and inferred immune-cell fractions. SSc: (A) gene–cell correlation network; (B) ridgeline plot of immune-cell fractions. APS: (C) gene–cell correlation network; (D) ridgeline plot of immune-cell fractions.

### miRNA network and candidate drug prediction

miRWalk prediction identified immune-regulatory miRNAs as potential upstream regulators of the shared biomarkers [24]. Network visualization highlighted miR-155-5p and miR-146a-5p as multi-target nodes (Fig 10; S5 Table), consistent with their established roles in inflammatory feedback and innate immune signaling [26,25].

**Fig 10.**
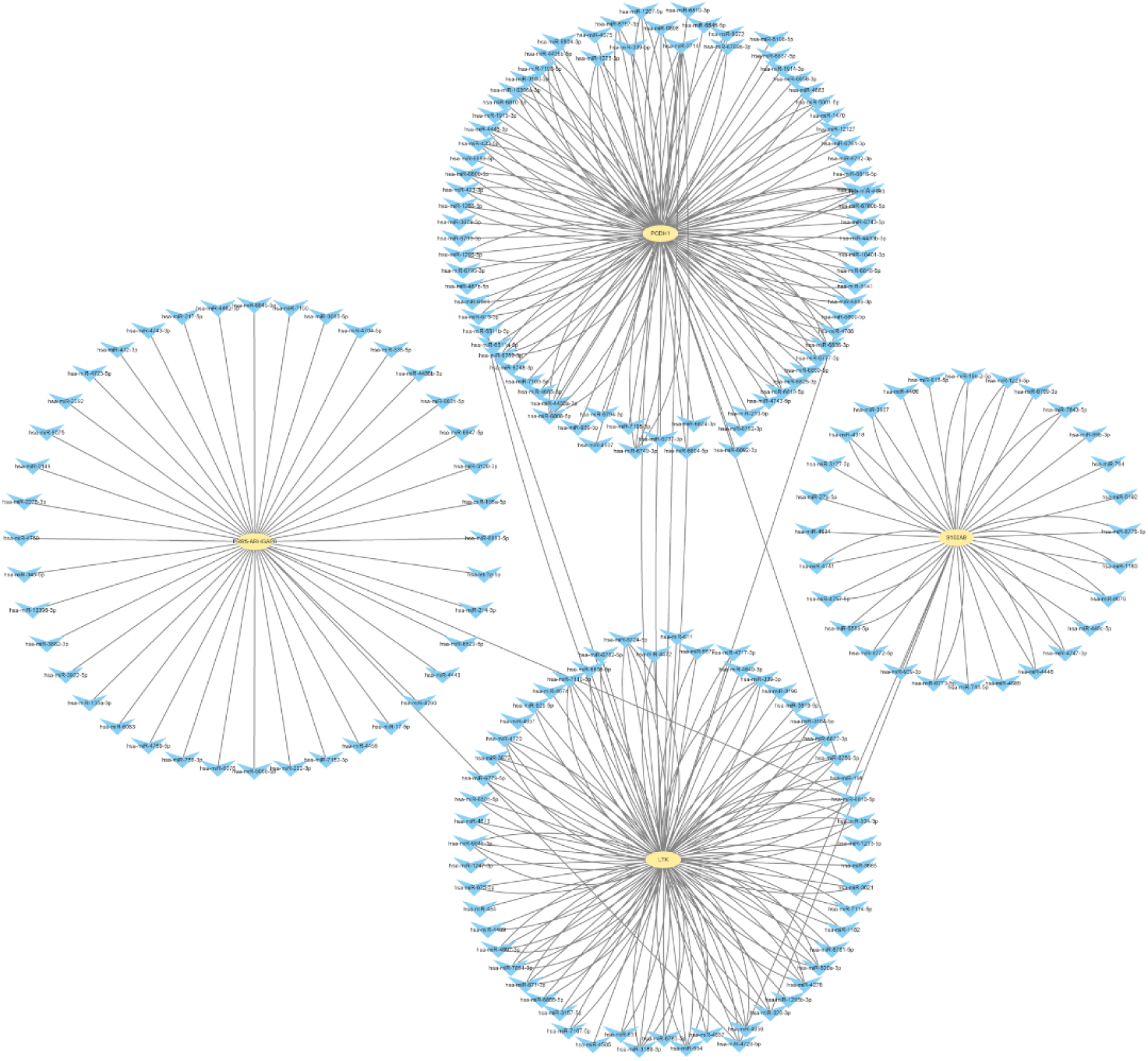
Predicted miRNA–mRNA regulatory network for shared biomarkers. miRWalk-predicted miRNAs targeting S100A8, LTK, PRR5-ARHGAP8, and PCDH1 are shown; IER5L-AS1 had no miRWalk targets in this analysis. Circles denote miRNAs and squares denote mRNAs; edges indicate predicted interactions. Notable multi-target miRNAs include miR-155-5p (targets S100A8 and LTK) and miR-146a-5p (targets S100A8 and PCDH1).

DSigDB enrichment nominated compounds whose transcriptional signatures overlapped with the biomarker genes, including celecoxib, tamibarotene, HMN-176, and XMD14-99 (Table 2; S6 Table) [27]. These results are intended for hypothesis generation and do not imply therapeutic efficacy in APS or SSc.

**Table 2.** Top candidate compounds from DSigDB enrichment analysis.

| Drug names | P-value | Adjusted P-value | Odds Ratio | Combined Score | Genes |
| --- | --- | --- | --- | --- | --- |
| XMD14-99<br>LINCS | 0.0057 | 0.0717 | 226.96 | 1171.31 | LTK |
| celecoxib<br>MCF7 | 0.0059 | 0.0717 | 217.08 | 1111.11 | S100A8 |
| Tamibarotene<br>CTD | 0.0073 | 0.0717 | 23.26 | 114.2303167 | PCDH1;S100A8 |
| HMN-176<br>CTD | 0.0077 | 0.0717 | 166.37 | 809.09 | PCDH1 |

## Discussion

### Principal findings

Across two peripheral blood transcriptome datasets, we identified a shared gene-expression program linking APS and SSc and distilled it to a five-gene candidate biomarker set using complementary machine-learning approaches. The 281 shared DEGs were enriched for immune and inflammatory pathways, including interferon-related programs, consistent with conserved transcriptional modules reported across autoimmune diseases [10]. The five prioritized genes (S100A8, IER5L-AS1, LTK, PRR5-ARHGAP8, and PCDH1) were selected independently in both cohorts and showed good within-cohort discrimination (AUC > 0.75) [22]. Immune deconvolution suggested that both diseases exhibit a myeloid-skewed blood profile [23], while SSc showed additional adaptive immune signals, in line with disease heterogeneity [4,5].

### Biological interpretation of prioritized biomarkers

S100A8 encodes a myeloid-associated calcium-binding protein highly expressed in neutrophils and monocytes and capable of amplifying inflammatory responses [31]. Its reproducible upregulation and correlation with inferred neutrophil fractions in both cohorts support a shared myeloid-inflammatory axis that may contribute to vascular injury and chronic inflammation in APS and SSc [32]. Prior work has linked the S100A8/A9 complex to Th17-related responses in autoimmune settings [32].

IER5L-AS1 is a long non-coding RNA (lncRNA) with limited functional characterization in APS and SSc. Because lncRNAs can modulate immune programs and contribute to autoimmune pathogenesis [33], its consistent dysregulation makes it a plausible regulatory candidate. Clarifying its cellular source and downstream targets will require targeted experimental work [33].

LTK is a receptor tyrosine kinase involved in cell signaling [34] and may reflect shared activation programs in circulating immune cells. PRR5-ARHGAP8 is linked to Rho GTPase-related signaling, and PCDH1 is involved in cell–cell adhesion and barrier-related functions [35]. One possibility is that these genes connect immune activation to endothelial dysfunction, tissue remodeling, and fibrotic processes; however, these are mechanistic hypotheses that require direct validation.

### Shared and disease-specific immune landscapes

The inferred immune landscapes showed both convergence and divergence. Both diseases exhibited increased neutrophil and monocyte/macrophage fractions, consistent with chronic inflammation and vascular involvement [15,23]. In APS, neutrophil extracellular traps have been implicated in thrombosis [15], whereas in SSc, macrophage polarization and T-cell imbalance contribute to fibrotic remodeling [5]. The additional increase in inferred CD4+ T-cell and NK-cell signals in SSc is consistent with broader adaptive immune dysregulation [5]. Overall, the gene–immune correlations suggest that the biomarker signals reflect immune-cell composition and/or activation states rather than purely cell-intrinsic transcriptional changes.

### Regulatory and translational hypotheses

miRNA prediction highlighted immune-regulatory miRNAs that may target multiple biomarkers [24]. Given their established roles in inflammatory feedback and innate immune signaling [26,25], these interactions provide concrete hypotheses for post-transcriptional regulation of the shared signature. For example, miR-155 inhibition can attenuate inflammation in autoimmune models [26], raising the possibility that it modulates components of the shared program in APS and SSc.

Drug-signature enrichment highlighted compounds including celecoxib, tamibarotene, HMN-176, and XMD14-99 [27]. These results simply indicate overlap between drug-induced expression signatures and the biomarker genes; they do not establish efficacy in APS or SSc. Follow-up experiments could test whether these compounds modulate the shared biomarkers or associated immune phenotypes in relevant models [28].

### Limitations and future directions

Several limitations merit emphasis. First, the APS cohort is small (9 cases), which reduces power and can inflate apparent performance [22]. Second, the datasets were generated on different microarray platforms; although we avoided direct merging and relied on concordant within-dataset results, residual platform effects may persist [19]. Third, this study is computational and based on public data, so independent cohort validation and mechanistic experiments are required [13]. Fourth, CIBERSORT provides relative estimates from bulk expression data [23]; orthogonal measurements such as flow cytometry or single-cell profiling would strengthen inference [17]. Finally, DSigDB results are intended for hypothesis generation and require experimental screening [28].

Future work should validate the five-gene panel in larger, multicenter cohorts and clarify the cellular sources of the shared signals using single-cell approaches [16]. Mechanistic studies focused on myeloid-inflammatory pathways (including S100A8-related signaling) may help connect the shared blood signature to disease-relevant immune and vascular phenotypes [31].

## Data Availability

All relevant data are within the manuscript and its Supporting Information files.

## Ethics statement

All data analyzed in this study were obtained from public GEO repositories. The original studies reported appropriate ethical approval and informed consent. This work involved secondary analysis of publicly available, de-identified data and did not include any new experiments involving human participants.

## Data availability

All transcriptome data are available from GEO under accession numbers GSE102215 and GSE231691. Code used for analysis is available from the corresponding author upon reasonable request.

## List of Abbreviations (Alphabetical Order)

ACR: American College of Rheumatology
ANN: artificial neural network
APS: antiphospholipid syndrome
AUC: area under the curve
CIBERSORT: cell-type identification by estimating relative subsets of RNA transcripts (CIBERSORT) algorithm
DEG: differentially expressed gene
DEGs: differentially expressed genes
DSigDB: Drug Signatures Database
EULAR: European Alliance of Associations for Rheumatology
FDR: false discovery rate
GEO: Gene Expression Omnibus
GO: Gene Ontology
GSEA: gene set enrichment analysis
KEGG: Kyoto Encyclopedia of Genes and Genomes
LM22: leukocyte gene signature matrix of 22 immune cell types
lncRNA: long non-coding RNA
miRNA: microRNA
mRNA: messenger RNA
NF-κB: nuclear factor kappa-light-chain-enhancer of activated B cells
NK: natural killer (cells)
RMA: robust multi-array average
ROC: receiver operating characteristic
SSc: systemic sclerosis
TNF: tumor necrosis factor

